# Indian Female Migrants Face Greater Barriers to Post-Covid Recovery than Males: Evidence from a Panel Study

**DOI:** 10.1101/2022.06.03.22275964

**Authors:** Jenna Allard, Maulik Jagnani, Yusuf Neggers, Rohini Pande, Simone Schaner, Charity Troyer Moore

**Affiliations:** University of Colorado Denver, Denver, CO, USA; University of Michigan; 735 S. State St. 4216, Ann Arbor, MI 48109, USA; Economic Growth Center, Yale University; 27 Hillhouse Avenue, New Haven, CT, 06511, USA; Center for Economic and Social Research, University of Southern California; 635 Downey Way, Los Angeles, CA 90089, USA; MacMillan Center, Yale University; 34 Hillhouse Avenue, New Haven, CT, 06511, USA

**Keywords:** domestic migrants, Covid-19 pandemic, panel, India, labor markets, food insecurity

## Abstract

**Background:** India’s abrupt nationwide Covid-19 lockdown internally displaced millions of urban migrants, who made arduous journeys to distant rural homes. Documenting their labor market reintegration is a critical aspect of understanding the economic costs of the pandemic for India’s poor. In a country marked by low and declining female labor force participation, identifying gender gaps in labor market reintegration – as a marker of both women’s vulnerability at times of crisis and setbacks in women’s agency – is especially important. Yet most studies of pandemic-displaced Indian migrants are small, rely on highly selected convenience samples, and lack a gender focus.

**Methods:** Beginning in April 2020 we enrolled roughly 4,600 displaced migrants who had returned to two of India’s poorest states into a panel survey, which tracked enrollees through July 2021. Survey respondents were randomly selected from the states’ official databases of return migrants, with sampling stratified by state and gender. 85 percent of enrollees (3,950) were working in urban areas prior to the pandemic. Our analysis focuses on a balanced panel of 1,780 workers who were interviewed three times through July 2021, considering labor market re-entry, earnings, and measures of vulnerability by gender.

**Findings:** Both men and women struggle to remigrate – by July 2021 (over a year after the nationwide lockdown ended), no more than 63 percent (95% CI [60,66]) of men and 55 percent [51,59] of women had left their home villages since returning. Initially, returning migrants transition from non-agricultural urban employment into agriculture and unemployment in rural areas. Alongside, incomes plummet, with both genders earning roughly 17 percent of their pre-lockdown incomes in July 2020. Remigration is critical to regaining income – male re-migrants report earnings on par with their pre-lockdown incomes by January 2021, while men remaining in rural areas earn only 23 percent [19,27] of their pre-pandemic income. Remigration benefits women to a lesser extent – female remigrants regain no more than 65 percent [57,73] of their pre-pandemic income at any point. This contrast reflects significantly higher rates of unemployment among women, both among those remaining in rural areas (9 percentage points [6,13] higher than men across waves) and among those who remigrate (13 percentage points [9,17] higher than men across waves). As a result, we observe gender gaps in well-being: female migrants were 7 percentage points [4,10] more likely to report reduced consumption of essential goods and fare 6 percentage points [4,7] worse on a food security index.

**Interpretation:** Return migrants of both genders experienced persistent hardships for over a year after the initial pandemic lockdown. Female migrants fare worse, driven by both lower rates of remigration and lower rates of labor market re-entry both inside and outside home villages. Some women drop out of the labor force entirely, but most unemployed report seeking or being available to work. In short, pandemic-induced labor market displacement has far-reaching, long-term consequences for migrant workers, especially women.

**Funding:** Survey costs were funded by research grants from IZA/FCDO Gender, Growth, and Labour Markets in Low Income Countries Programme, J-PAL Jobs and Opportunity Initiative, and the Evidence-based Measures of Empowerment for Research on Gender Equality (EMERGE) program at University of California San Diego. Funders had no role in study design, study implementation, data analysis, or manuscript preparation.

**Research in context:** *Evidence before this study:* Most research documenting the experience of displaced domestic migrants during the pandemic is focused on difficulties faced in returning to their home villages and the immediate consequences of this displacement. Existing evidence has found high levels of short-run economic and psychological distress, especially among women and children, and under-coverage of government programs designed to ease the lockdown’s sudden economic shock.

*Added value of this study:* This study contributes to existing literature by surveying a large sample of male and female workers, designed to be broadly representative of returned migrants in two of India’s poorest states. Our work takes a longer-term view, tracking study participants’ efforts to remigrate and reintegrate into the labor force over 15 months. We document sustained difficulties attaining pre-pandemic levels of income and consumption insecurity, especially among women, who struggle even after remigrating.

*Implications of all the available evidence:* Taken as a whole, the evidence underscores that displaced Indian migrants are a vulnerable and underserved social group, who have faced (and will likely continue to face) lasting negative effects of the Covid-19 pandemic. Displaced migrants – and especially women – would likely benefit from programs designed to facilitate re-entry into urban labor markets; wrap around services that address other effects of the pandemic (e.g. psychological distress) may be particularly valuable.

## 1 Introduction

On March 24, 2020, India responded to the Covid-19 pandemic by announcing one of the largest and strictest lockdowns in the world. Announced with just four hours notice, the nationwide lockdown caused millions of low-income migrant workers to return to home villages, some traveling on foot and others via crowded specially-designated buses and trains. Once returned, migrants were often placed in government-run quarantine centers, facing stigma and uncertain job prospects. The economic consequences were particularly severe for these migrants, families dependent on remittances, and other informal workers.^2;3;10;11;17;30;39^ The nationwide lockdown was lifted on May 31, 2020, and the country began opening up in stages until March 2021, when the Delta variant devastated the country. New state-managed lockdowns were announced, threatening a fragile economic recovery, and precipitating another (smaller-scale) return of migrants to rural areas.^6;9;40^

Domestic rural-urban migration is how many households in lower-income countries exit poverty,^5;14;15;16;36^ and it is also an important contributor to economic growth.^35;38^ While female migration in India is largely linked to marriage, the number of women migrating for work had been increasing rapidly pre-pandemic.^42^ Weak re-integration of displaced migrants into urban labor markets thus both threatens household well-being and economic growth.

Adverse impacts of displacement may be especially salient for female migrants: Indian women are some of the least employed in the world, with only 18.6 percent in the labor force per the nationally-representative 2018-19 Periodic Labor Force Survey (PLFS), and rural women’s labor force participation has declined over the past twenty years, in part due to lack of jobs considered suitable for women.^20^ Yet research shows that women’s access to higher-paying formal sector jobs – such as those more widely available in urban areas – increases women’s career aspirations while delaying marriage and child bearing,^31^ which is, in turn, associated with better outcomes for women and their families.^19;26;41^ Thus, Covid-induced job dislocation and urban displacement could have lasting negative implications for a female migrant’s agency and her family’s well-being.

More broadly, women around the world have felt the impact of pandemic-induced challenges disproportionately,^4;7;27;37;43^ and evidence suggests India is no exception. The pandemic has differentially reduced Indian women’s employment,^1;24;25^ imposed additional childcare responsibilities,^21^ and increased rates of mental distress and domestic violence among the general female population.^12^

While researchers have sought to understand vulnerable migrants’ well-being and the challenges they face through the pandemic,^8;13;18;22;23;32;33;34^ most studies have been short term, lack a gender focus, and use convenience samples – such as stranded migrants seeking assistance from NGOs – that may be especially prone to selection bias.

This study contributes to the existing literature by documenting longer-term experiences of migrant workers, with a focus on gender gaps in remigration, labor market integration, and economic hardship. We leverage unique partnerships with two of India’s poorest states, which granted our research team access to databases with information on over 435,000 migrant workers who returned to their home villages in the immediate aftermath of India’s first lockdown. We follow economically active urban migrants displaced by the nationwide lockdown for a 15 month period, tracking their short and longer-term ability to recover, and their resilience through a second wave of lockdown restrictions and health threats.

## 2 Methods

### 2.1 Study design

Our panel study includes four survey rounds conducted between April 2020 and June 2021 with a sample of urban migrants who returned during the national lockdown to their home states of Bihar and Chhattisgarh in north and central India, respectively.^1^ Sample respondents were drawn from databases of migrants registered as returnees by government officials between April and June 2020. In Bihar, government officials collected contact information from all returning migrants at inter-state transport hubs, yielding a list of approximately 651,000 individuals, which was accessed by our research team in April and May 2020. In Chhattisgarh, local officials collected contact information from returning migrants when they arrived at their designated local quarantine center, yielding a list of 580,968 migrants, which was accessed by our research team in May and June 2020.^2^ Before selecting individuals to be surveyed, we dropped those under the age of 18 from these lists and inferred gender based on name.

### 2.2 Survey procedure

We administered phone-based surveys of return migrants across four rounds: April to June 2020; July to August 2020; January to March 2021; and June to July 2021. Survey rounds roughly corresponded to the initial nationwide lockdown, two periods of relative recovery, and then the tail end of India’s Delta-variant Covid surge. For the first survey round, we interviewed 5,879 individuals randomly chosen from the then-current return migrant lists, stratifying to achieve equal proportions by state-gender category.^3^ To increase the likelihood of survey completion, we attempted to reach selected individuals on up to six different dates, and contacted individuals were given the option to reschedule surveys for a more convenient time and date. If we were unable to reach a selected respondent, we randomly chose a replacement in the same state-gender category. Surveys were conducted by trained enumerators and took approximately 30 minutes.

In survey rounds two through four, we targeted completion of at least 1,000 surveys per gender in each state. To create state-by-gender calling lists in each round, we prioritized respondents that had completed surveys in previous rounds, then respondents with failed contact attempts or incomplete surveys from previous rounds, and finally individuals who had not yet been contacted – randomly ordering individuals within each group. If in a given round we were unable to reach a selected respondent, a replacement was chosen from the same state-gender ordered list. We completed surveys with 4,644 individuals in round two, 4,829 in round three, and 4,318 in round four.^4^ Overall, 45 percent of respondents interviewed in round two were also surveyed in rounds three and four. Within these balanced-panel respondents, 99 percent of men and 67 percent of women were working before the pandemic.^5^

We restrict our analysis sample to respondents who were employed immediately prior to the initial nationwide lockdown. To track individual transitions in and out of work over time, we limit our attention to migrant workers surveyed in rounds two through four (round one, which was conducted when many migrants were still in quarantine centers, was focused on immediate distress and pre-pandemic outcomes). Together these restrictions yield a sample of 1,780 migrant workers, 1,202 male and 578 female.^6^

### 2.3 Outcomes

We focus on a set of core indicators relevant to migrant well-being throughout the pandemic. The first four relate to work and economic activities and include whether the migrant reported working for pay in the week prior to the survey; whether the migrant had re-migrated (versus remaining in one’s home village); earnings in the previous week; and that same earnings value as a share of the migrant’s pre-pandemic weekly earnings.^7^ The second set of indicators focuses on migrant consumption, notably whether the migrant reported their household reducing essential consumption in the month prior to the survey and a food insecurity index for the previous week – defined as the share of affirmative answers to three questions based on indicators from the Food Insecurity Experience Scale,^28^ with one taken from each of the low, moderate, and severe ranges of the scale.^8^

### 2.4 Empirical approach

Before presenting main results, we compare our sample of migrant workers to a representative sample of workers in Bihar and Chhattisgarh from the 2017-19 Periodic Labour Force Survey (PLFS). The PLFS comes with person-level weights designed to be representative of the population. Since our main analysis is unweighted with roughly equal numbers of migrants from Bihar and Chhattisgarh, we further weight our PLFS subsample to put equal weight on each state.

Our analysis focuses on the migrants panel described above, where we use unpaired t-tests allowing for unequal variances to compare means across male and female respondents.

## 3 Results

### 3.1 Migrant characteristics pre-pandemic

Table 1 compares our balanced panel sample of return migrants to a representative sample of Bihari and Chhattisgarhi workers in the PLFS. Means are presented separately for men (columns 1 and 4) and women (columns 2 and 5). The differences in means, 95 percent confidence intervals, and the p-values from t-tests of the equality of means across genders for unpaired data with unequal variances are given in columns 3 and 6.

**Table 1:**
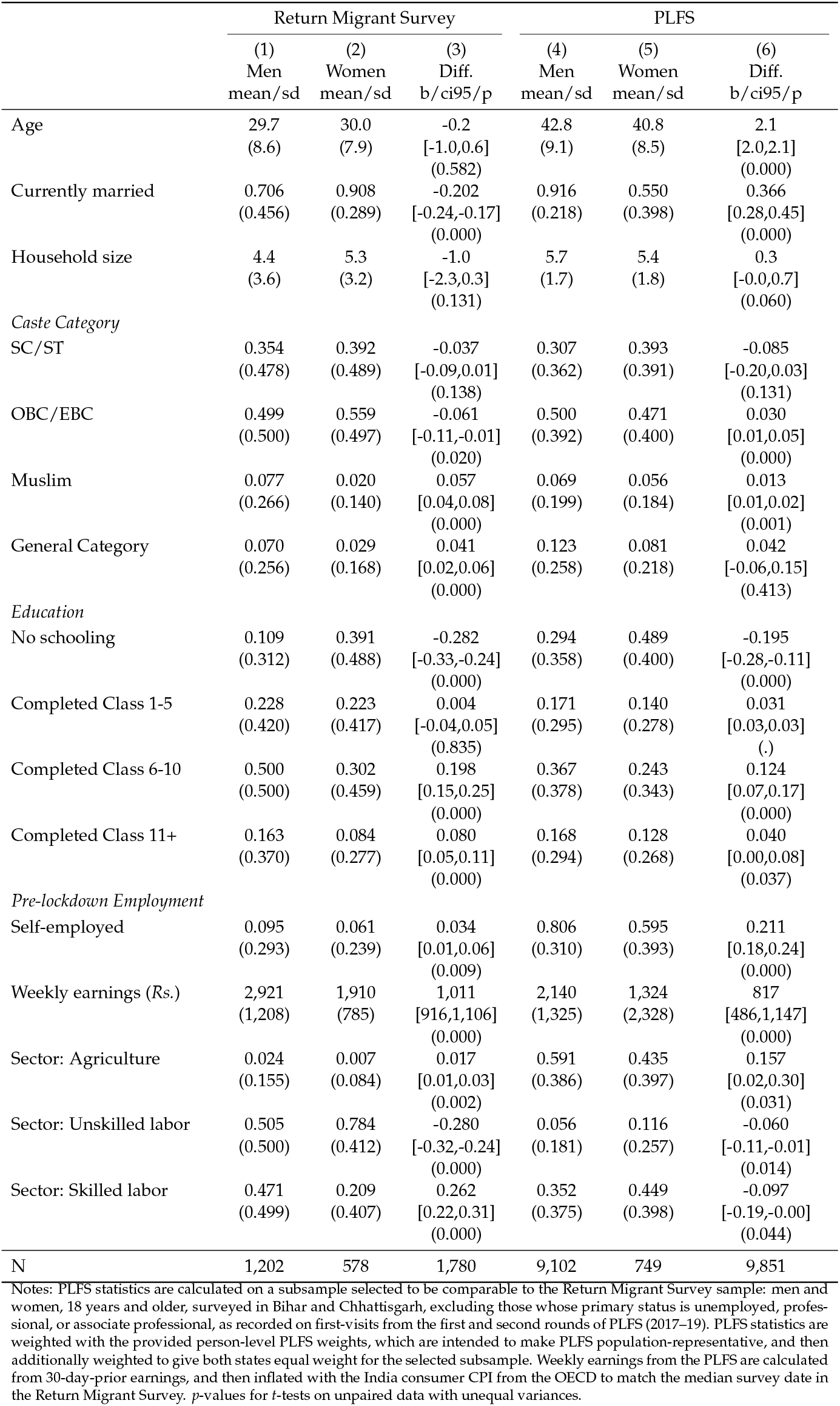
Sample pre-pandemic characteristics

Our migrants are 10 to 13 years younger than the average state-based worker, with male migrants less likely to be married than the general working population and female migrants more likely to be married. The high marriage rates observed for female migrant workers, but not female workers based in Bihar and Chhattisgarh, likely reflect prevailing gender norms regarding work and marriage: Women, especially those with young children, are often expected to stay home and take care of the household. Alongside, migration for work may reduce women’s marital prospects as their chastity is called into question; therefore, women may be more likely to migrate once married.^9^

More than 85 percent of the migrants sample belongs to officially recognized marginalized groups as members of “Scheduled Caste” (SC), “Scheduled Tribe” (ST), or “Other or Extremely Backward Class” (OBC/EBC) groups; only 7 percent of male migrants and 3 percent of female migrants belong to the least disadvantaged “General Category” caste groups. Though still low, rates of general category caste memberships are higher in the state-based workers sample, reflecting disproportionate social marginalization among the migrant sample.

Yet the migrant sample is better educated than the general population of workers from Bihar and Chhattisgarh, with 11 percent of male migrants and 39 percent of female migrants reporting no schooling, compared to 29 and 49 percent of men and women in the state-based sample. We also observe notable differences in terms of pre-lockdown employment: less than 10 percent of migrant workers were self-employed, compared to 81 percent of men and 60 percent of women from the PLFS sample. While a significant share of state-based workers are employed in the agricultural sector (59 percent of men and 44 percent of women), most migrants work outside agriculture in either unskilled (51 percent of men and 78 percent of women) or skilled (47 percent of men and 21 percent of women) labor. Finally, male and female migrants reported pre-lockdown weekly earnings of Rs 2,921 and Rs 1,910 respectively.^10^ In line with research documenting high returns to domestic migration, these incomes are substantially higher than statewide means of Rs 2,140 and Rs 1,324 among men and women respectively.

To summarize, migrant workers are more likely to belong to disadvantaged social groups, but are better educated and earn more. Female migrants fare significantly worse than their male peers in terms of most measures of advantage, underscoring their vulnerability.

### 3.2 How did the pandemic and lockdown impact migrant outcomes?

#### Remigration

We begin our analysis of migrant workers’ post-lockdown trajectories by examining remigration by gender in Figure 1. Remigration took time: 90 percent of men and 96 percent of women were in their home villages during recovery phase 1 (July 2020), more than a month after the nationwide lockdown ended. By recovery phase 2 (January 2021), 53 percent of men and 46 percent of women were outside their home villages. Yet just 37 percent of men and 35 percent of women remained outside their home village as of Covid wave 2 (June 2021), reflecting a new round of migrant returns that coincided with the Delta variant wave. Looking across waves, 63 percent of men remigrated at least once, compared to 55 percent of women (Diff: 0.08, 95% CI [0.03,0.13]). This difference suggests women faced greater barriers to remigration than men, which could have important consequences for their labor market and socioeconomic outcomes.

**Figure 1:**
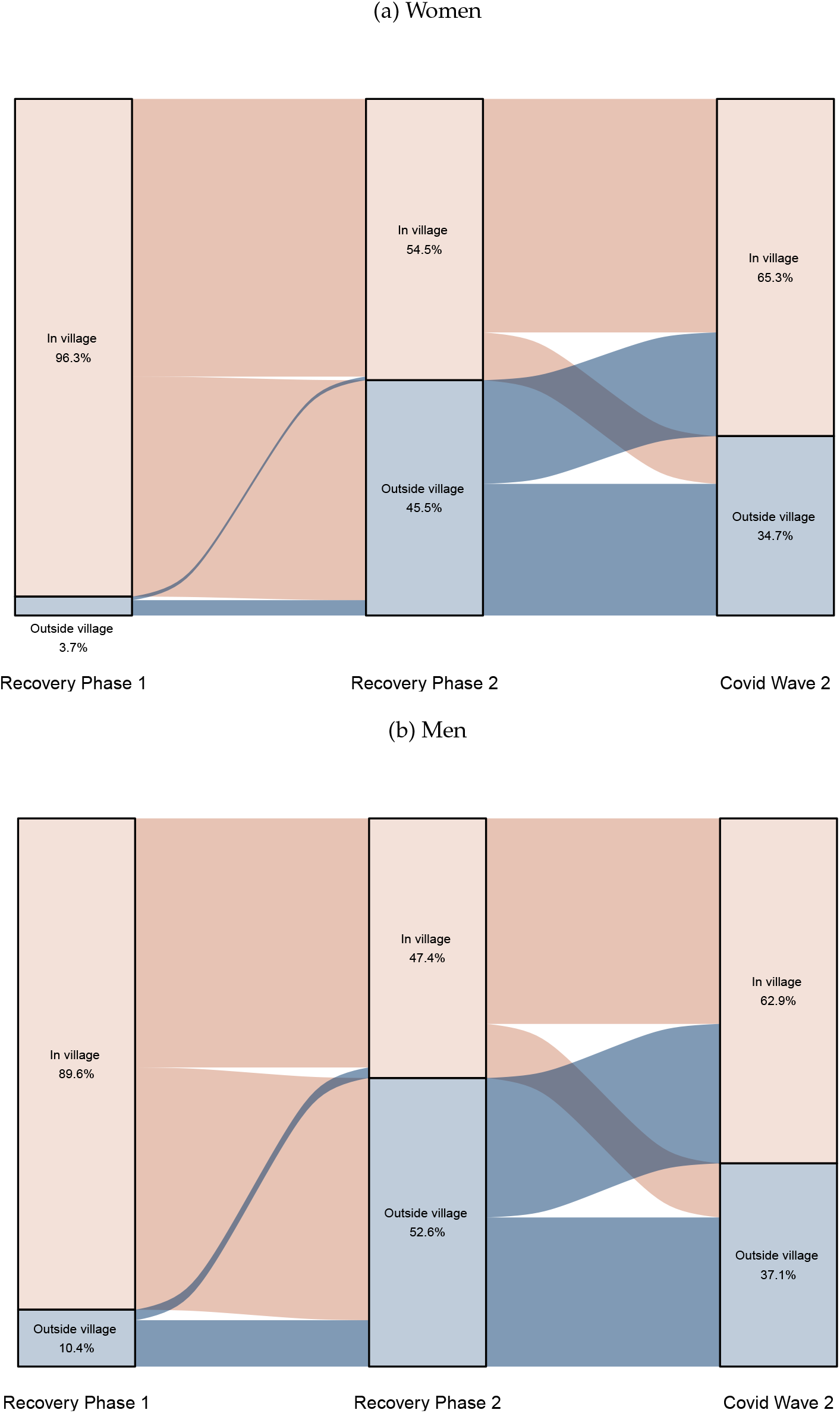
Remigration status - by gender

#### Employment

Figures 2 and 3 consider migrant transitions in and out of employment sectors. Figure 2 shows that following the lockdown, the majority of unskilled workers – both male and female – transitioned into agricultural work, while larger shares of skilled workers transitioned into unemployment. While unemployment rates in the immediate aftermath of the lockdown are similar for both genders (36 to 40 percent), unemployment among women steadily grows, first to 42 percent during recovery phase 2 and then 53 percent during Covid wave 2. Men, in contrast, are more likely to find work over the longer term – just 25 and 37 percent of men are unemployed at recovery phase 2 and Covid wave 2, respectively. This reflects two phenomena: women who transition into unemployment are significantly less likely to transition back out in the following round (women: 41 percent, men: 63 percent; Diff: -22, 95% CI [-27,-16]) *and* women who are working in a given round are significantly more likely to exit into unemployment than their male peers (women: 40 percent, men: 28 percent; Diff: 12, 95% CI [8,16]). Higher rates of female unemployment do not simply reflect lower rates of remigration, as Figure 3 shows that women are more likely than men to be unemployed even conditional on remigration status.

**Figure 2:**
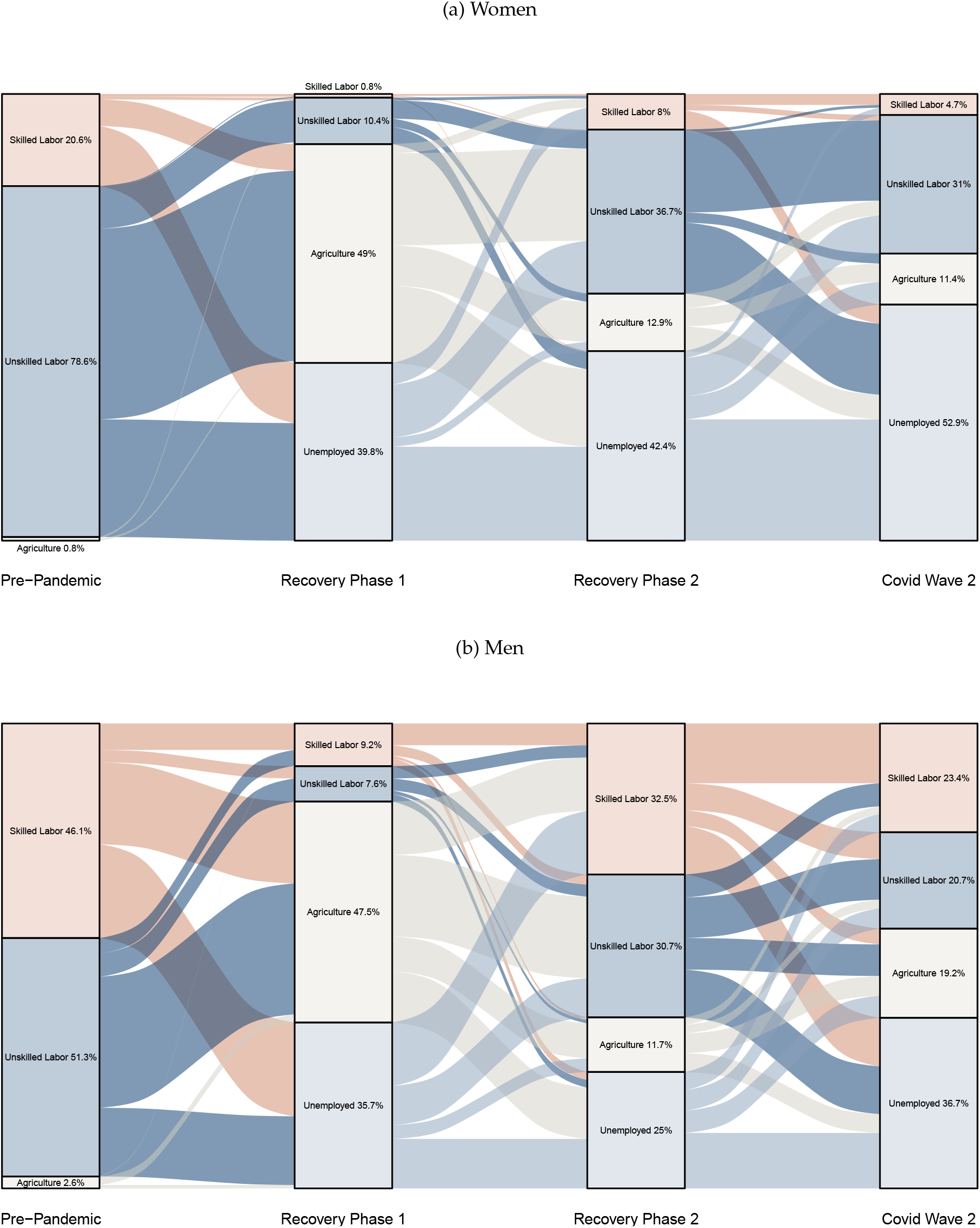
Sector of employment - by gender

**Figure 3:**
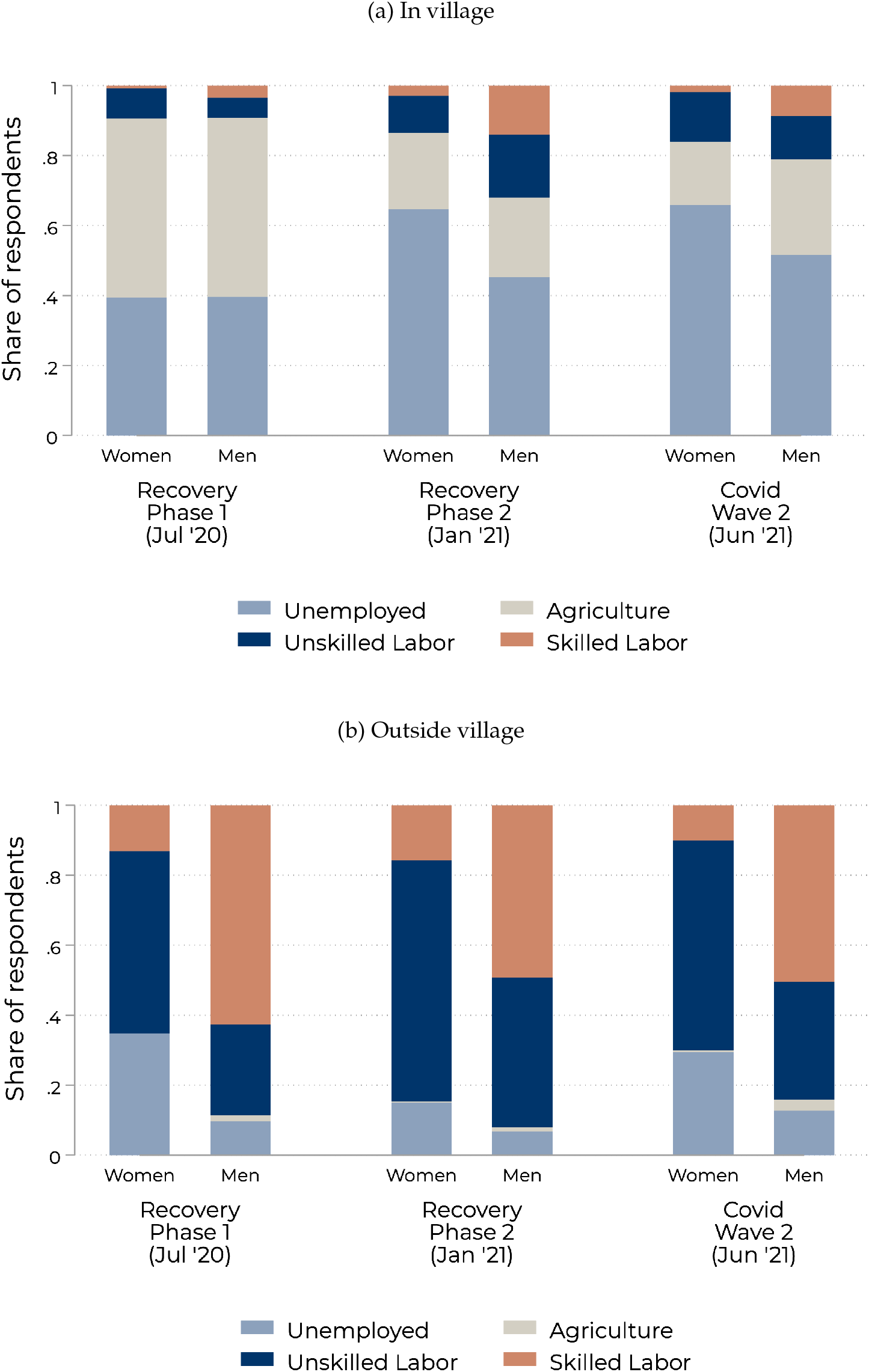
Sector of employment - by remigration status

The unemployed category includes both those actively seeking or ready to work and those no longer seeking work. Though large majorities of both unemployed men and women report seeking or being willing to accept a job across survey rounds, women are significantly less likely than men to do so (women: 86 percent, men: 94 percent; Diff: -8, 95% CI [-11,-5]). Among unemployed and not seeking work, 60 percent of women state the reason as due to “domestic duties or pregnancy”, compared to 27 percent of men (Diff: 33, 95% CI [19,48]). 18 percent of women indicate “wages too low or no work available”, compared to 6 percent of men (Diff: 12, 95% CI [3,21]). In contrast, only 15 percent of women state “health problems or fear of COVID”, relative to 42 percent of men (Diff: -27, 95% CI [-40,-13]).

Beyond falling into unemployment, Figures 2 and 3 show that women who do stay in the labor market struggle to access high-return work. While men also face challenges finding skilled jobs – with a post-pandemic share in skilled labor peaking in recovery phase 2 at 33 percent (or 70 percent of pre-pandemic prevalence) – a maximum of 8 percent of women (also in recovery phase 2; 39 percent of pre-pandemic prevalence) are observed working in skilled labor post-pandemic (Diff: -24, 95% CI [-27,-20]).

#### Earnings

Panel (a) of Figure 4 presents results relevant to understanding how the above-described labor market transitions relate to income. Recall from Figures 1 and 2 that in recovery phase 1, most migrants remained in their home villages, working in agriculture. As a result, incomes plummeted, with both men and women earning under Rs 500 (USD 7) per week on average, less than 20 percent of their pre-lockdown income. Moreover, the gender earnings gap shrank to just Rs 151 (95% CI [41,260]), reflecting poor labor market opportunities for men and women alike. As the recovery progressed, average male income rebounded significantly, reaching Rs 1,670 by January 2021, or 64 percent of pre-pandemic levels. Women, on the other hand, continued to struggle, with average earnings reaching just 35 percent of pre-pandemic levels by recovery phase 2. The resulting gender gap of Rs 1,036 is statistically significant (95% CI [896,1,177]). While the Delta variant wave induced a second round of wage compression, men continued to outpace women in June 2021, earning Rs 1,111 per week on average compared to women’s weekly earnings of Rs 490 (Diff: Rs 620, 95% CI [497,744]), or 40 percent of pre-lockdown earnings for men as compared to 28 percent for women (Diff: 12, 95% CI [7,18]).

**Figure 4:**
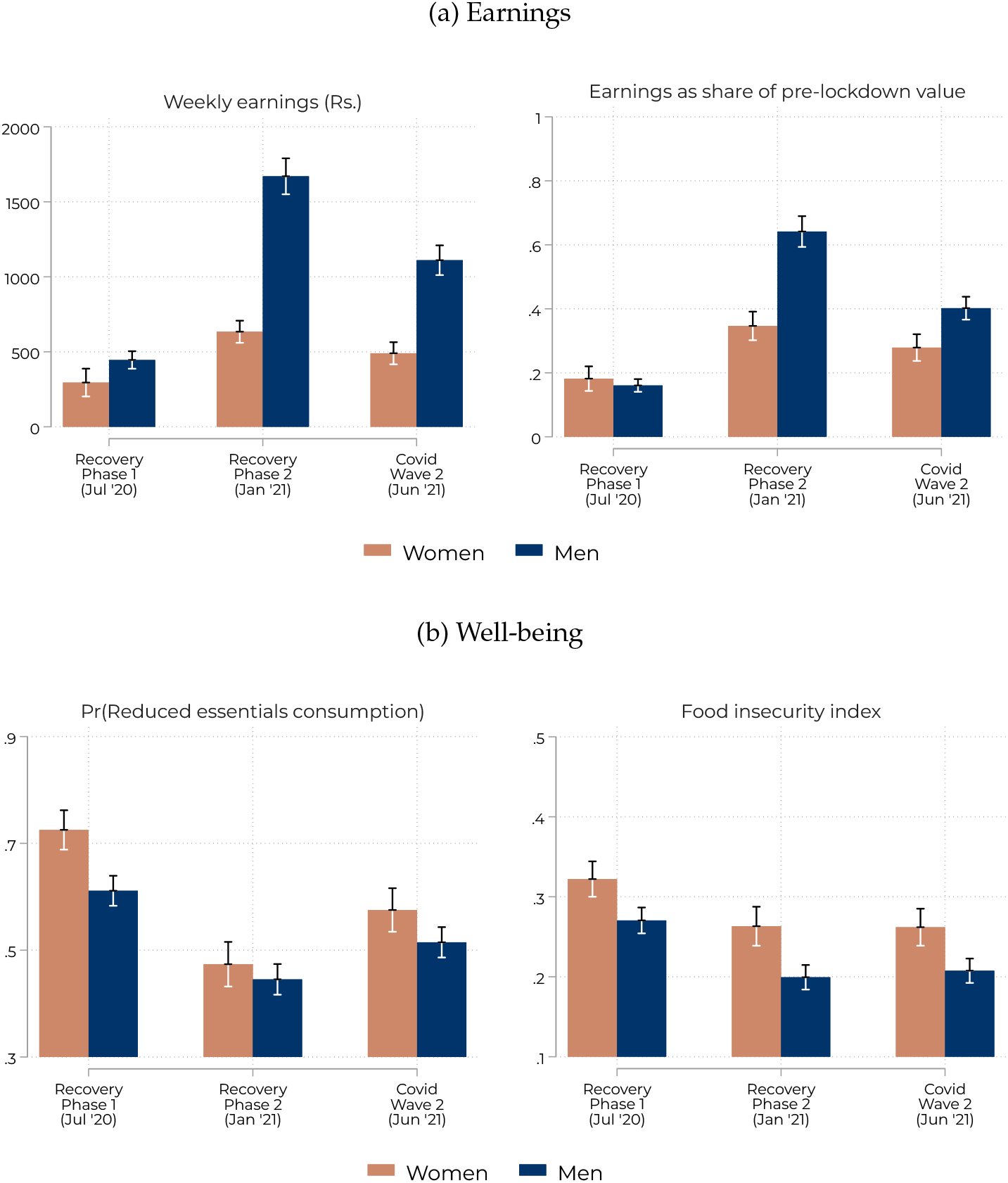
Earnings and well-being

#### Well-being

Panel (b) of Figure 4 points to the implications of these economic gender gaps. First, women are significantly more vulnerable: pooling across waves, women are 7 percentage points more likely to report cutting back on essentials in their household (95% CI [4,10]) with an average 0.06 unit higher food insecurity index score (95% CI [0.04,0.07]). Second, although rates of distress were highest in recovery phase 1, when most migrants were still in their home villages earning little income, they remained persistently high across survey rounds: in recovery phase 2, for example, roughly 45 percent of both men and women reported reducing essential consumption to make ends meet.^11^

### 3.3 Implications of remigration for earnings and well-being

Figure 5 asks whether differences in remigration can explain the observed gender differences in earnings and well-being. Panel (a) graphs earnings by gender and migration status. Overall earnings levels – and gender gaps – among those in their home villages remain low. Men in their home villages earn on average just 10 to 23 percent of their pre-pandemic income, and women earn only 11 to 17 percent. The story is very different for those who remigrate. First, though it takes time, men who remigrate recover much of their lost income. Average weekly income among male remigrants peaks at Rs 2,665 (USD 38) in recovery phase 2, amounting to a full recovery of pre-pandemic income for the average male remigrant. Women, however, continue to struggle: average female earnings also peak in recovery phase 2, but at a significantly lower Rs 1,200 (Diff: Rs 1,465, 95% CI [1,270,1,660]), or 65 percent of pre-pandemic levels.

**Figure 5:**
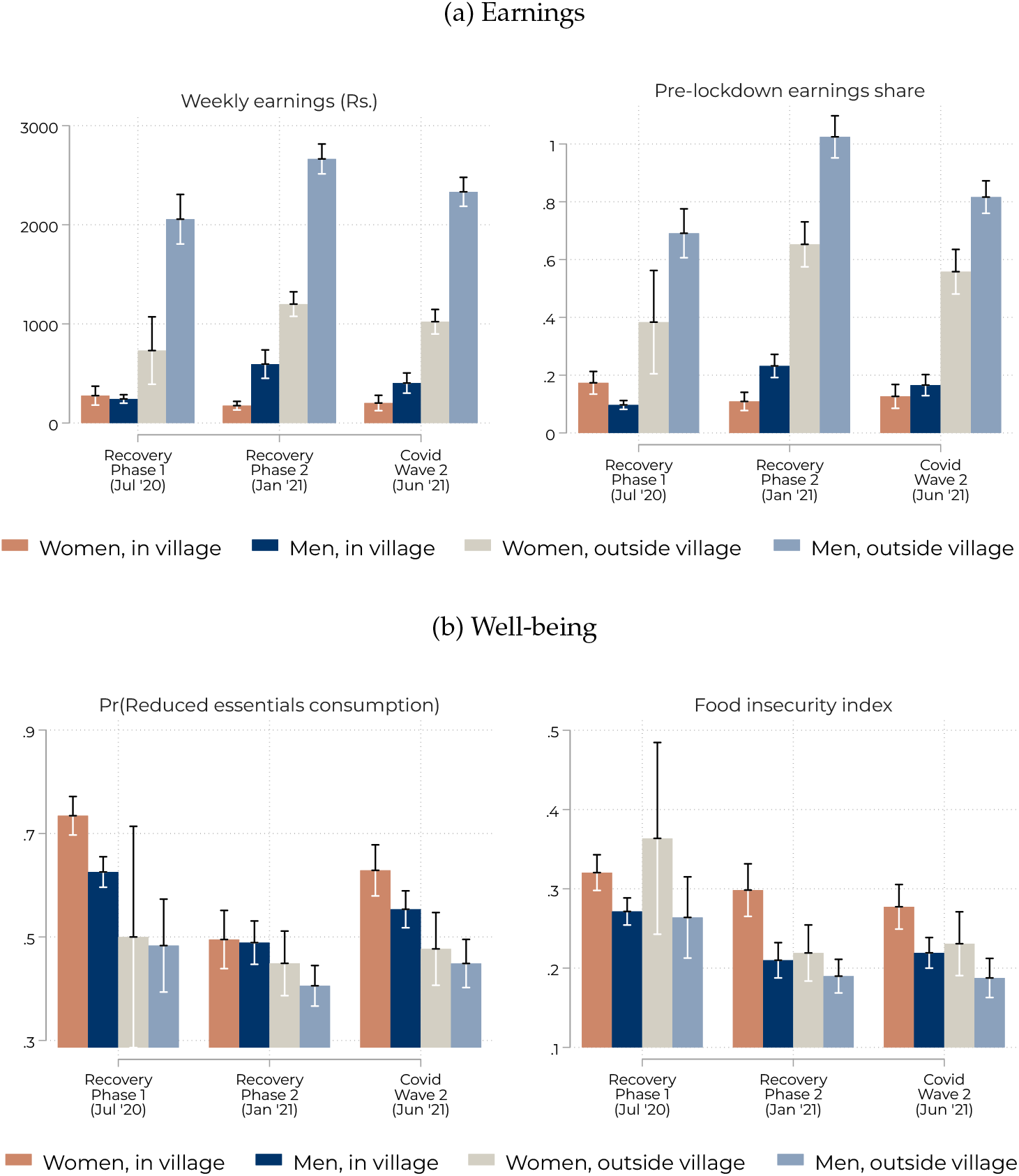
Earnings and well-being - by remigration status

Panel (b) of Figure 5 plots markers of distress by gender and remigration status. In general, both men and women who re-migrate are less likely to report cutting back consumption and have lower food insecurity index scores.^12^ Overall, our analysis suggests that women faced greater challenges re-integrating into the labor market than their male peers, even conditional on managing to re-migrate. These gender differences are large and persistent, and could have significant implications for women’s long-run labor market prospects.

## 4 Discussion

Rural to urban migration is a pathway out of poverty for many households, particularly as agricultural employment opportunities diminish as economies industrialize and develop. For Indian women, in particular, rural to urban migration offers an important opportunity to connect to higher-paying jobs rarely available to them in rural areas. Against this background, we show that the Covid-19 pandemic had adverse long-term repercussions for displaced urban migrants, exacerbating already-large gender gaps in economic outcomes.

Fifteen months after the pandemic began, displaced migrants earned significantly less and were likely to be working in lower-skilled jobs than they had been previously, with female earnings and skilled job losses larger than those of men. Remigration emerged as an important income recovery strategy for men, but less so for women – on average male remigrants were able to recover all of their pre-pandemic income prior to the Delta surge, while female remigrants continued to struggle, experiencing lower income and higher unemployment. As a consequence, women remained persistently more vulnerable (as measured by reduced consumption and food security) throughout our entire study period.

This paper’s contribution is twofold – to track the evolution of economic outcomes through the pandemic with a large sample of displaced migrants returning to low-income states in rural India, and to quantify emerging gender gaps in economic reintegration. Our work does have some limitations. First, our research design does not allow us to estimate the causal effect of the pandemic, nor does it allow us to identify the underlying mechanisms that hamper women’s recovery relative to men’s. While our sampling strategy was designed to be representative of large pool of returned migrant workers, surveys were conducted over the phone, meaning our study under-represents the experiences of those who lacked phone numbers or regular access to a phone. Finally, our analysis sample is limited to a subset of individuals who participated in the last three waves of our survey. While survey non-response could relate to migrant economic activity and well-being, our results are stable across a variety of sample restrictions, painting a consistent picture of elevated distress and significant gender gaps in the aftermath of India’s nationwide lockdown.

Our research raises concerns about the pandemic’s longer-term consequences for women, especially related to their ability to access meaningful work via urban labor markets. In many ways, our sample of migrants includes some of the most intrepid women in India’s work force, willing to transgress social norms and brave safety concerns in order to secure more remunerative work outside their home villages. Their post-pandemic struggles point to an urgent need to better understand the barriers faced by female job seekers and identify policies that facilitate women’s access to high-return jobs.

## Data Availability

De-identified datasets used for the analysis presented in this manuscript and code will be posted on an online open data platform prior to final publication. Until that point, data is available upon reasonable request from Jenna Allard.

## Contributors

All authors contributed equally to this paper. All authors had full access to the data.

## Declaration of interests

The authors declare that they have no competing interests.

## Acknowledgments

We are grateful to Akash Bhatt, Nils Enevoldsen, Akshita Mehra, Vyana McIntyre, Urvi Naik, Abhishek Sharma, Tejita Tiwari, and Tanya Vaidya for excellent research and management assistance throughout the data collection and analysis phases. Survey costs were funded by research grants from Institute of Labour Economics (IZA)/UK Aid (FCDO) Gender, Growth, and Labour Markets in Low Income Countries Programme (GA-5-700), J-PAL Jobs and Opportunity Initiative (JOI-1374), and the Evidence-based Measures of Empowerment for Research on Gender Equality (EMERGE) program at University of California San Diego (INV-018007). EMERGE and its director, Anita Raj, facilitated submission of the manuscript as part of a series on Covid and gender. Other than this, the funders had no role in the writing or editing of the manuscript, the decision to submit the manuscript for publication, or any other aspects relevant to this manuscript. All authors accept responsibility to submit for publication.

## Ethics approval and consent to participate

Research protocols were approved through the Yale Human Research Protection Program for protocol #2000027920, and for the IFMR review board (in India, for Inclusion Economics India Centre at IFMR, who collected the data) for protocol #IRB00007107. All survey respondents consented to participate in the study in line with research protocols.

## 5 Appendix

**Table 1:** Sample pre-pandemic characteristics: panel respondents v. attritors

|  | Men |  |  | Women |  |  |
| --- | --- | --- | --- | --- | --- | --- |
|  | (1)<br>Panel<br>mean/sd | (2)<br>Attritors<br>mean/sd | (3)<br>Diff.<br>b/ci95/p | (4)<br>Panel<br>mean/sd | (5)<br>Attritors<br>mean/sd | (6)<br>Diff.<br>b/ci95/p |
| Age | 29.7<br>(8.6) | 29.1<br>(9.3) | -0.6<br>[-1.3,0.1]<br>(0.076) | 30.0<br>(7.9) | 29.4<br>(8.4) | -0.5<br>[-1.4,0.3]<br>(0.224) |
| Currently married | 0.706<br>(0.456) | 0.643<br>(0.479) | -0.064<br>[-0.10,-0.03]<br>(0.001) | 0.908<br>(0.289) | 0.839<br>(0.368) | -0.070<br>[-0.10,-0.03]<br>(0.000) |
| Household size | 5.1<br>(3.2) | 5.0<br>(3.1) | -0.0<br>[-0.3,0.2]<br>(0.746) | 5.2<br>(2.9) | 5.2<br>(3.2) | 0.0<br>[-0.3,0.4]<br>(0.831) |
| *Caste Category* |  |  |  |  |  |  |
| SC/ST | 0.354<br>(0.478) | 0.436<br>(0.496) | 0.082<br>[0.04,0.12]<br>(0.000) | 0.392<br>(0.489) | 0.468<br>(0.499) | 0.076<br>[0.02,0.13]<br>(0.006) |
| OBC/EBC | 0.499<br>(0.500) | 0.416<br>(0.493) | -0.082<br>[-0.12,-0.04]<br>(0.000) | 0.559<br>(0.497) | 0.472<br>(0.500) | -0.088<br>[-0.14,-0.03]<br>(0.002) |
| Muslim | 0.077<br>(0.266) | 0.071<br>(0.258) | -0.005<br>[-0.03,0.02]<br>(0.624) | 0.020<br>(0.140) | 0.028<br>(0.164) | 0.008<br>[-0.01,0.02]<br>(0.362) |
| General Category | 0.070<br>(0.256) | 0.076<br>(0.265) | 0.006<br>[-0.02,0.03]<br>(0.600) | 0.029<br>(0.168) | 0.033<br>(0.179) | 0.004<br>[-0.02,0.02]<br>(0.689) |
| *Education* |  |  |  |  |  |  |
| No schooling | 0.109<br>(0.312) | 0.156<br>(0.363) | 0.048<br>[0.02,0.07]<br>(0.000) | 0.391<br>(0.488) | 0.380<br>(0.486) | -0.011<br>[-0.06,0.04]<br>(0.684) |
| Completed Class 1-5 | 0.228<br>(0.420) | 0.205<br>(0.404) | -0.023<br>[-0.05,0.01]<br>(0.160) | 0.223<br>(0.417) | 0.210<br>(0.407) | -0.014<br>[-0.06,0.03]<br>(0.543) |
| Completed Class 6-10 | 0.500<br>(0.500) | 0.475<br>(0.500) | -0.025<br>[-0.06,0.01]<br>(0.208) | 0.302<br>(0.459) | 0.287<br>(0.452) | -0.015<br>[-0.06,0.03]<br>(0.541) |
| Completed Class 11+ | 0.163<br>(0.370) | 0.164<br>(0.370) | 0.000<br>[-0.03,0.03]<br>(0.978) | 0.084<br>(0.277) | 0.124<br>(0.329) | 0.040<br>[0.01,0.07]<br>(0.015) |
| *Pre-lockdown Employment* |  |  |  |  |  |  |
| Self-employed | 0.095<br>(0.293) | 0.086<br>(0.280) | -0.009<br>[-0.03,0.01]<br>(0.430) | 0.061<br>(0.239) | 0.070<br>(0.255) | 0.009<br>[-0.02,0.04]<br>(0.482) |
| Weekly earnings (Rs.) | 2,921<br>(1,208) | 2,934<br>(1,258) | 14<br>[-84,111]<br>(0.784) | 1,910<br>(785) | 1,840<br>(780) | -70<br>[-158,18]<br>(0.122) |
| Sector: Agriculture | 0.024<br>(0.155) | 0.027<br>(0.162) | 0.003<br>[-0.01,0.02]<br>(0.684) | 0.007<br>(0.084) | 0.019<br>(0.138) | 0.012<br>[-0.00,0.02]<br>(0.054) |
| Sector: Unskilled labor | 0.505<br>(0.500) | 0.529<br>(0.499) | 0.024<br>[-0.02,0.06]<br>(0.226) | 0.784<br>(0.412) | 0.755<br>(0.430) | -0.029<br>[-0.08,0.02]<br>(0.225) |
| Sector: Skilled labor | 0.471<br>(0.499) | 0.444<br>(0.497) | -0.027<br>[-0.07,0.01]<br>(0.179) | 0.209<br>(0.407) | 0.226<br>(0.418) | 0.017<br>[-0.03,0.06]<br>(0.475) |
| N | 1,202 | 1,374 | 2,576 | 578 | 800 | 1,378 |

Notes: p-values for t-tests on unpaired data with unequal variances.

**Table 2:** Sample characteristics by wave and gender

|  | Recovery Phase 1 (Jul '20) |  |  | Recovery Phase 2 (Jan '21) |  |  | Covid Wave 2 (Jun '21) |  |  |
| --- | --- | --- | --- | --- | --- | --- | --- | --- | --- |
|  | (1)<br>Men<br>mean/sd | (2)<br>Women<br>mean/sd | (3)<br>Diff.<br>b/ci95/p | (4)<br>Men<br>mean/sd | (5)<br>Women<br>mean/sd | (6)<br>Diff.<br>b/ci95/p | (7)<br>Men<br>mean/sd | (8)<br>Women<br>mean/sd | (9)<br>Diff.<br>b/ci95/p |
| Outside village | 0.103<br>(0.305) | 0.040<br>(0.196) | 0.063<br>[0.04,0.09]<br>(0.000) | 0.525<br>(0.500) | 0.456<br>(0.498) | 0.070<br>[0.02,0.12]<br>(0.006) | 0.370<br>(0.483) | 0.348<br>(0.477) | 0.022<br>[-0.03,0.07]<br>(0.366) |
| Sector: Skilled labor | 0.096<br>(0.294) | 0.012<br>(0.110) | 0.084<br>[0.06,0.10]<br>(0.000) | 0.324<br>(0.468) | 0.087<br>(0.281) | 0.237<br>[0.20,0.27]<br>(0.000) | 0.241<br>(0.428) | 0.047<br>(0.212) | 0.194<br>[0.16,0.22]<br>(0.000) |
| Sector: Unskilled labor | 0.079<br>(0.270) | 0.105<br>(0.306) | -0.026<br>[-0.06,0.00]<br>(0.085) | 0.308<br>(0.462) | 0.367<br>(0.483) | -0.059<br>[-0.11,-0.01]<br>(0.015) | 0.203<br>(0.402) | 0.298<br>(0.458) | -0.096<br>[-0.14,-0.05]<br>(0.000) |
| Sector: Agriculture | 0.460<br>(0.499) | 0.490<br>(0.500) | -0.030<br>[-0.08,0.02]<br>(0.235) | 0.116<br>(0.320) | 0.122<br>(0.327) | -0.006<br>[-0.04,0.03]<br>(0.716) | 0.184<br>(0.388) | 0.122<br>(0.328) | 0.062<br>[0.03,0.10]<br>(0.000) |
| Unemployed | 0.360<br>(0.480) | 0.390<br>(0.488) | -0.030<br>[-0.08,0.02]<br>(0.225) | 0.246<br>(0.431) | 0.419<br>(0.494) | -0.172<br>[-0.22,-0.13]<br>(0.000) | 0.373<br>(0.484) | 0.537<br>(0.499) | -0.164<br>[-0.21,-0.11]<br>(0.000) |
| Weekly earnings (Rs.) | 446<br>(968) | 295<br>(1,117) | 151<br>[41,260]<br>(0.007) | 1,670<br>(2,091) | 634<br>(887) | 1,036<br>[896,1,177]<br>(0.000) | 1,111<br>(1,737) | 490<br>(892) | 620<br>[497,744]<br>(0.000) |
| Earnings as share of pre-lockdown value | 0.160<br>(0.325) | 0.182<br>(0.453) | -0.021<br>[-0.06,0.02]<br>(0.334) | 0.642<br>(0.831) | 0.346<br>(0.526) | 0.295<br>[0.23,0.36]<br>(0.000) | 0.402<br>(0.621) | 0.279<br>(0.496) | 0.123<br>[0.07,0.18]<br>(0.000) |
| Reduced consumption of essentials | 0.611<br>(0.488) | 0.725<br>(0.447) | -0.114<br>[-0.16,-0.07]<br>(0.000) | 0.445<br>(0.497) | 0.474<br>(0.500) | -0.028<br>[-0.08,0.02]<br>(0.273) | 0.515<br>(0.500) | 0.575<br>(0.495) | -0.060<br>[-0.11,-0.01]<br>(0.017) |
| Food security index | 0.270<br>(0.281) | 0.322<br>(0.268) | -0.052<br>[-0.08,-0.02]<br>(0.000) | 0.199<br>(0.266) | 0.263<br>(0.292) | -0.064<br>[-0.09,-0.03]<br>(0.000) | 0.208<br>(0.268) | 0.262<br>(0.280) | -0.054<br>[-0.08,-0.03]<br>(0.000) |
| N | 1,201 | 577 | 1,778 | 1,201 | 578 | 1,779 | 1,198 | 576 | 1,774 |

Notes: p-values for t-tests on unpaired data with unequal variances.

**Table 3:**
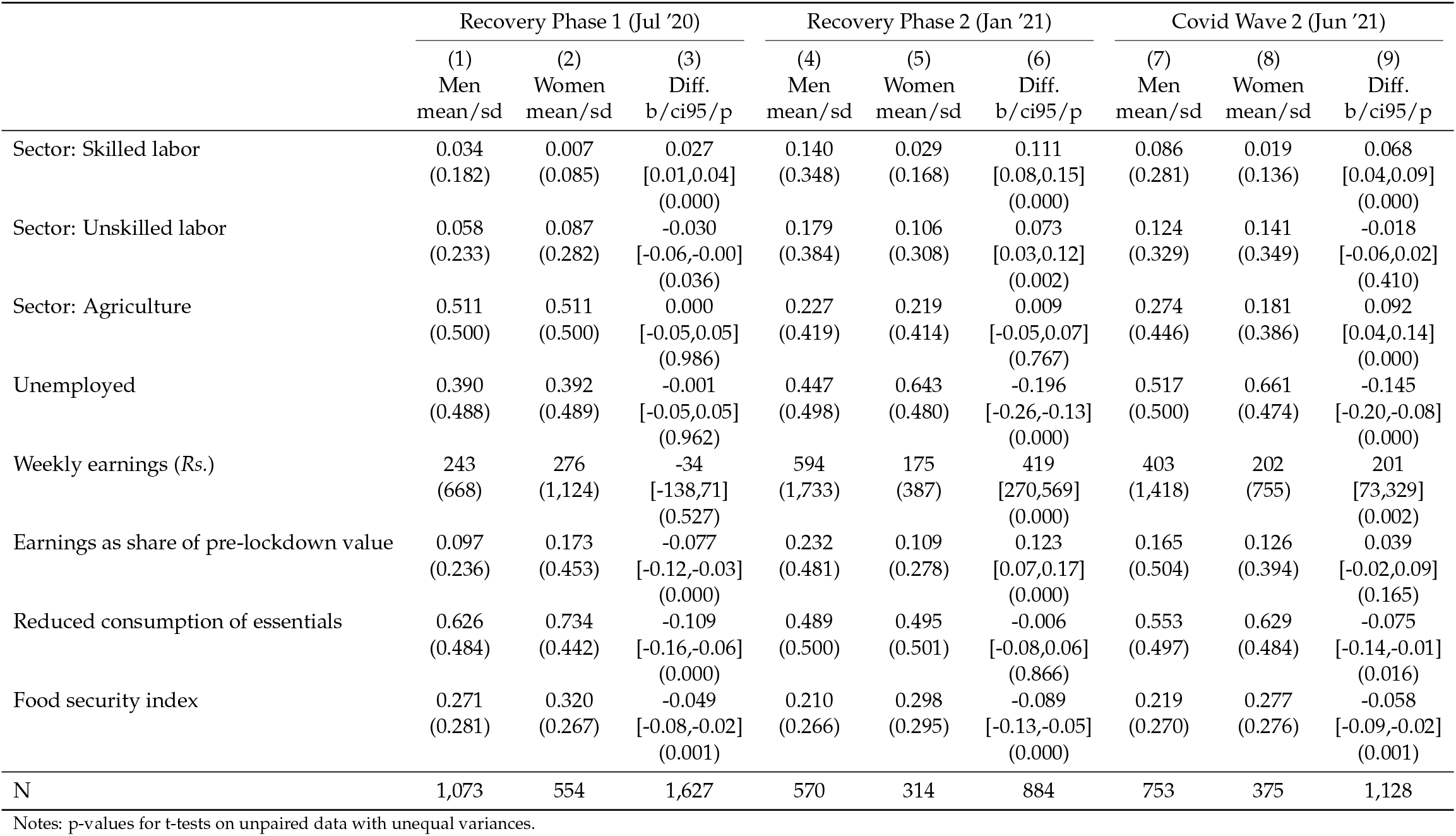
Sample characteristics by wave and gender, inside village only

|  | Recovery Phase 1 (Jul '20) |  |  | Recovery Phase 2 (Jan '21) |  |  | Covid Wave 2 (Jun '21) |  |  |
| --- | --- | --- | --- | --- | --- | --- | --- | --- | --- |
|  | (1)<br>Men<br>mean/sd | (2)<br>Women<br>mean/sd | (3)<br>Diff.<br>b/ci95/p | (4)<br>Men<br>mean/sd | (5)<br>Women<br>mean/sd | (6)<br>Diff.<br>b/ci95/p | (7)<br>Men<br>mean/sd | (8)<br>Women<br>mean/sd | (9)<br>Diff.<br>b/ci95/p |
| Sector: Skilled labor | 0.034<br>(0.182) | 0.007<br>(0.085) | 0.027<br>[0.01,0.04]<br>(0.000) | 0.140<br>(0.348) | 0.029<br>(0.168) | 0.111<br>[0.08,0.15]<br>(0.000) | 0.086<br>(0.281) | 0.019<br>(0.136) | 0.068<br>[0.04,0.09]<br>(0.000) |
| Sector: Unskilled labor | 0.058<br>(0.233) | 0.087<br>(0.282) | -0.030<br>[-0.06,-0.00]<br>(0.036) | 0.179<br>(0.384) | 0.106<br>(0.308) | 0.073<br>[0.03,0.12]<br>(0.002) | 0.124<br>(0.329) | 0.141<br>(0.349) | -0.018<br>[-0.06,0.02]<br>(0.410) |
| Sector: Agriculture | 0.511<br>(0.500) | 0.511<br>(0.500) | 0.000<br>[-0.05,0.05]<br>(0.986) | 0.227<br>(0.419) | 0.219<br>(0.414) | 0.009<br>[-0.05,0.07]<br>(0.767) | 0.274<br>(0.446) | 0.181<br>(0.386) | 0.092<br>[0.04,0.14]<br>(0.000) |
| Unemployed | 0.390<br>(0.488) | 0.392<br>(0.489) | -0.001<br>[-0.05,0.05]<br>(0.962) | 0.447<br>(0.498) | 0.643<br>(0.480) | -0.196<br>[-0.26,-0.13]<br>(0.000) | 0.517<br>(0.500) | 0.661<br>(0.474) | -0.145<br>[-0.20,-0.08]<br>(0.000) |
| Weekly earnings (Rs.) | 243<br>(668) | 276<br>(1,124) | -34<br>[-138,71]<br>(0.527) | 594<br>(1,733) | 175<br>(387) | 419<br>[270,569]<br>(0.000) | 403<br>(1,418) | 202<br>(755) | 201<br>[73,329]<br>(0.002) |
| Earnings as share of pre-lockdown value | 0.097<br>(0.236) | 0.173<br>(0.453) | -0.077<br>[-0.12,-0.03]<br>(0.000) | 0.232<br>(0.481) | 0.109<br>(0.278) | 0.123<br>[0.07,0.17]<br>(0.000) | 0.165<br>(0.504) | 0.126<br>(0.394) | 0.039<br>[-0.02,0.09]<br>(0.165) |
| Reduced consumption of essentials | 0.626<br>(0.484) | 0.734<br>(0.442) | -0.109<br>[-0.16,-0.06]<br>(0.000) | 0.489<br>(0.500) | 0.495<br>(0.501) | -0.006<br>[-0.08,0.06]<br>(0.866) | 0.553<br>(0.497) | 0.629<br>(0.484) | -0.075<br>[-0.14,-0.01]<br>(0.016) |
| Food security index | 0.271<br>(0.281) | 0.320<br>(0.267) | -0.049<br>[-0.08,-0.02]<br>(0.001) | 0.210<br>(0.266) | 0.298<br>(0.295) | -0.089<br>[-0.13,-0.05]<br>(0.000) | 0.219<br>(0.270) | 0.277<br>(0.276) | -0.058<br>[-0.09,-0.02]<br>(0.001) |
| N | 1,073 | 554 | 1,627 | 570 | 314 | 884 | 753 | 375 | 1,128 |

Notes: p-values for t-tests on unpaired data with unequal variances.

**Table 4:**
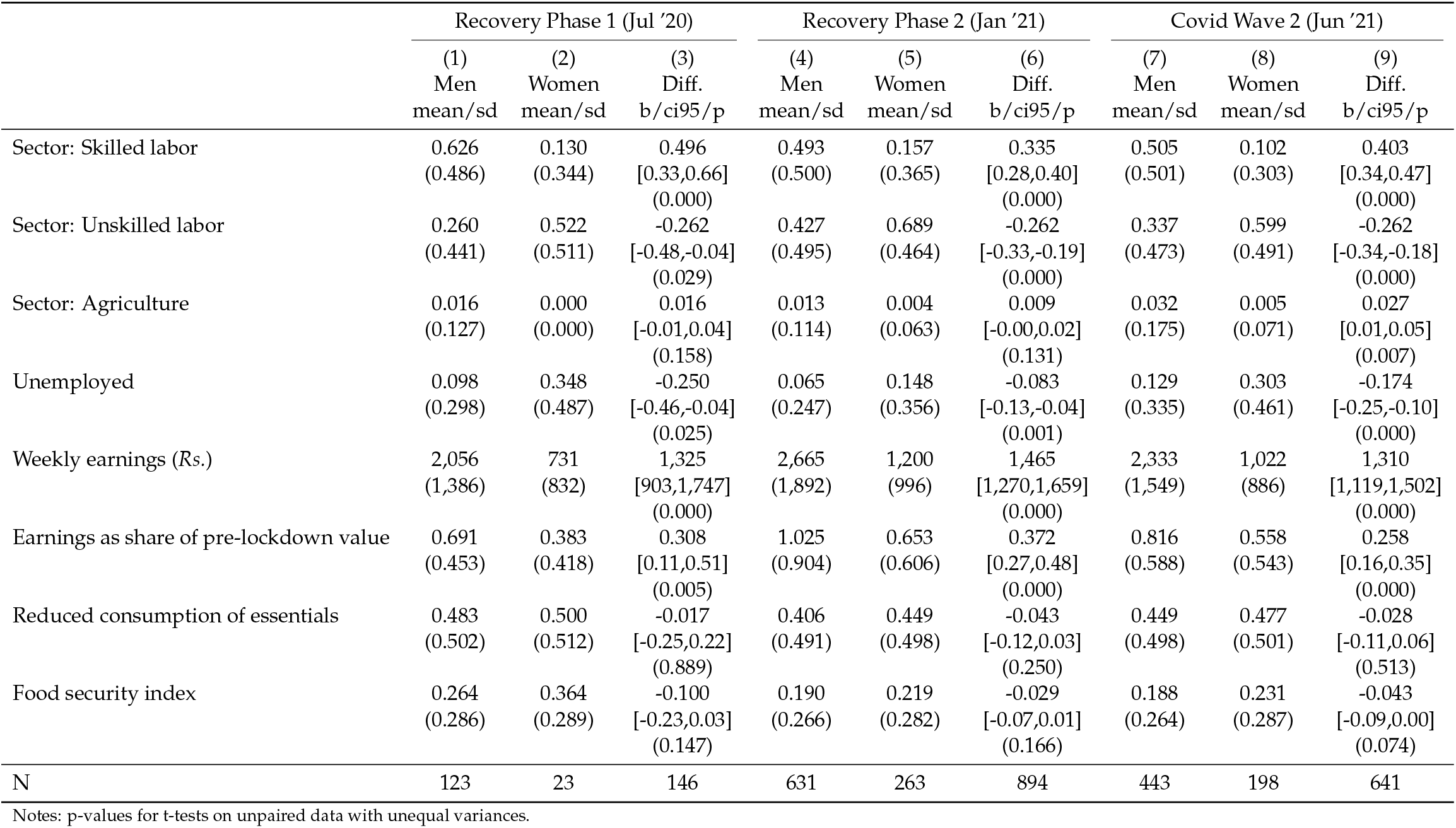
Sample characteristics by wave and gender, outside village only

|  | Recovery Phase 1 (Jul '20) |  |  | Recovery Phase 2 (Jan '21) |  |  | Covid Wave 2 (Jun '21) |  |  |
| --- | --- | --- | --- | --- | --- | --- | --- | --- | --- |
|  | (1)<br>Men<br>mean/sd | (2)<br>Women<br>mean/sd | (3)<br>Diff.<br>b/ci95/p | (4)<br>Men<br>mean/sd | (5)<br>Women<br>mean/sd | (6)<br>Diff.<br>b/ci95/p | (7)<br>Men<br>mean/sd | (8)<br>Women<br>mean/sd | (9)<br>Diff.<br>b/ci95/p |
| Sector: Skilled labor | 0.626<br>(0.486) | 0.130<br>(0.344) | 0.496<br>[0.33,0.66]<br>(0.000) | 0.493<br>(0.500) | 0.157<br>(0.365) | 0.335<br>[0.28,0.40]<br>(0.000) | 0.505<br>(0.501) | 0.102<br>(0.303) | 0.403<br>[0.34,0.47]<br>(0.000) |
| Sector: Unskilled labor | 0.260<br>(0.441) | 0.522<br>(0.511) | -0.262<br>[-0.48,-0.04]<br>(0.029) | 0.427<br>(0.495) | 0.689<br>(0.464) | -0.262<br>[-0.33,-0.19]<br>(0.000) | 0.337<br>(0.473) | 0.599<br>(0.491) | -0.262<br>[-0.34,-0.18]<br>(0.000) |
| Sector: Agriculture | 0.016<br>(0.127) | 0.000<br>(0.000) | 0.016<br>[-0.01,0.04]<br>(0.158) | 0.013<br>(0.114) | 0.004<br>(0.063) | 0.009<br>[-0.00,0.02]<br>(0.131) | 0.032<br>(0.175) | 0.005<br>(0.071) | 0.027<br>[0.01,0.05]<br>(0.007) |
| Unemployed | 0.098<br>(0.298) | 0.348<br>(0.487) | -0.250<br>[-0.46,-0.04]<br>(0.025) | 0.065<br>(0.247) | 0.148<br>(0.356) | -0.083<br>[-0.13,-0.04]<br>(0.001) | 0.129<br>(0.335) | 0.303<br>(0.461) | -0.174<br>[-0.25,-0.10]<br>(0.000) |
| Weekly earnings (Rs.) | 2,056<br>(1,386) | 731<br>(832) | 1,325<br>[903,1,747]<br>(0.000) | 2,665<br>(1,892) | 1,200<br>(996) | 1,465<br>[1,270,1,659]<br>(0.000) | 2,333<br>(1,549) | 1,022<br>(886) | 1,310<br>[1,119,1,502]<br>(0.000) |
| Earnings as share of pre-lockdown value | 0.691<br>(0.453) | 0.383<br>(0.418) | 0.308<br>[0.11,0.51]<br>(0.005) | 1.025<br>(0.904) | 0.653<br>(0.606) | 0.372<br>[0.27,0.48]<br>(0.000) | 0.816<br>(0.588) | 0.558<br>(0.543) | 0.258<br>[0.16,0.35]<br>(0.000) |
| Reduced consumption of essentials | 0.483<br>(0.502) | 0.500<br>(0.512) | -0.017<br>[-0.25,0.22]<br>(0.889) | 0.406<br>(0.491) | 0.449<br>(0.498) | -0.043<br>[-0.12,0.03]<br>(0.250) | 0.449<br>(0.498) | 0.477<br>(0.501) | -0.028<br>[-0.11,0.06]<br>(0.513) |
| Food security index | 0.264<br>(0.286) | 0.364<br>(0.289) | -0.100<br>[-0.23,0.03]<br>(0.147) | 0.190<br>(0.266) | 0.219<br>(0.282) | -0.029<br>[-0.07,0.01]<br>(0.166) | 0.188<br>(0.264) | 0.231<br>(0.287) | -0.043<br>[-0.09,0.00]<br>(0.074) |
| N | 123 | 23 | 146 | 631 | 263 | 894 | 443 | 198 | 641 |

Notes: p-values for t-tests on unpaired data with unequal variances.

## Footnotes

1 Both Bihar (pop. 100 million) and Chhattisgarh (pop. 25 million) are low-income states. However, per the 2011 Census, Bihar is a high out-migration state, with roughly 8 million Biharis living outside the state, while Chhattsigarh is a relatively low out-migration state, with roughly 0.5 million out-migrants.^29^

2 We designed these registration forms in collaboration with state government officials and a local non-profit organization. The registration portal was open from May 29, 2020 to June 25, 2020.

3 In Chhattisgarh, sampling was also stratified by the state’s five divisions, which are made up of groups of districts. In some cases, the total number of women in a stratum was less than the desired sample, so all women were included.

4 Small incentives of Rs 50 or 100 in phone credit were provided to respondents for survey completion in rounds two through four. Since return migrants were often under significant economic stress, enumerators in these rounds also gave respondents the option to receive information about resources to help them locate jobs and additional support.

5 This difference likely reflects in large part the importance of marriage as a reason for female out-of-state migration, with women accompanying their migrant worker husbands.

6 While Appendix Table 1 shows significant differences in some pre-pandemic characteristics between our analysis sample and attritors, our findings are essentially unchanged when the subsequent analysis is conducted using the full set of respondents in rounds two through four (results available upon request).

7 We elicited pre-pandemic earnings in intervals. When calculating earnings as a share of pre-pandemic earnings, we assign each respondent a pre-pandemic earnings value equal to the arithmetic mean of the upper and lower bounds of their earnings interval. The earnings intervals are Rs 1-5,000, 5-10,000, 10-15,000, 15-20,000, 20-25,000, and 25-35,000.

8 The three indicators are for experiencing the following in the previous week due to lack of resources: “worry that you will run out of food”, “ate less than you normally would”, and “went an entire day without eating”.

9 Most unmarried female workers in the PLFS data are widowed or divorced; thus never-married women account for a very small share of both migrants and state-based workers.

10 USD 41 and 27 using a January 2020 exchange rate of Rs 71 per USD

11 One limitation of our analysis is that we lack data on pre-pandemic levels of distress and therefore cannot measure how post-pandemic incidence of consumption cutbacks and food insecurity compares to pre-pandemic values.

12 The one exception is that female re-migrants have a higher food insecurity index score in Recovery Phase 1, though this difference is not statistically significant, likely reflecting the very low remigration rates we observe in this round of the survey.

## Notes

### Competing Interest Statement

The authors have declared no competing interest.

### Author Declarations

The Yale Human Research Protection Program of Yale University gave ethical approval for this work. The IFMR Human Subjects Committee of the Institute for Financial Management and Research gave ethical approval for this work.

